# Risk of apnoea-related cardiorespiratory instability in preterm infants is modulated by clinical, demographic and dynamic indicators

**DOI:** 10.64898/2026.05.13.26353101

**Authors:** Yiru Chen, Vithushanan Ketheeswaranathan, Surina Fordington, Luke Baxter, Freya Stevens, Coen S. Zandvoort, Rose Gawthorpe, Mauricio Villarroel, Luc Berthouze, Caroline Hartley

## Abstract

**Background:** Apnoea of prematurity is common and may cause desaturation and/or bradycardia. There is marked variability in infants’ cardiorespiratory responses to apnoea, despite standardised clinical thresholds. Factors influencing apnoea-related cardiorespiratory instability and whether instability can be predicted warrant investigation.

**Methods:** 181,511 apnoeas >5 seconds were identified from continuous physiological recordings from 146 preterm infants <37 weeks’ postmenstrual age. Cardiorespiratory instability was defined as bradycardia (>30% heart rate reduction) and/or oxygen desaturation (<85%). Mixed-effects models assessed clinical, demographic and dynamic modulators of the relationship between apnoea duration and cardiorespiratory instability. Machine learning (XGBoost) was used to train models to predict apnoea-related cardiorespiratory instability.

**Results:** Longer duration apnoeas were associated with increased instability, although variability was substantial and 3.6% of apnoeas <10 seconds were associated with cardiorespiratory instability, while 61.2% of apnoeas ≥20 seconds were not. Multiple clinical/demographic (postmenstrual and gestational age, sex, weight z-score, and ventilation mode) and dynamic (baseline heart rate, oxygen saturation, and recent apnoea clustering) factors were associated with increased instability risk. Apnoea-related cardiorespiratory instability could be predicted with a balanced test accuracy of 75.8% when incorporating all features, while a model using only clinical/demographic features achieved 66.0%.

**Conclusions:** Multiple factors influence cardiorespiratory responses to apnoea. Predictive modelling may enable personalised apnoea definitions, improving individualised care.

Impact statement

**Impact statement:** **What is the key message of your article?**

- We investigated variability in cardiorespiratory instability following apnoea in preterm infants.

**What does it add to the existing literature?**

- We demonstrate multiple factors which influence the cardiorespiratory changes following apnoea and develop a machine learning model which can accurately predict apnoea-related cardiorespiratory instability.

**What is the impact?**

- Prediction of cardiorespiratory instability could enable personalised apnoea alarms and treatment.

## 1. Introduction

Apnoea of prematurity is a potentially life-threatening condition that affects more than half of preterm-born infants^1^. Apnoea – the cessation of breathing – can lead to substantial hypoxia and bradycardia, as well as changes in cerebral oxygenation and suppression of brain activity^2–4^. Moreover, there may be long-term effects of apnoea, including increased risk of retinopathy of prematurity and neurodevelopmental impairments, likely driven by hypoxaemia^5,6^. Since apnoea occurrence is closely linked to gestational age at birth, it can be challenging to tease apart the long-term impacts of apnoea itself from the complications of prematurity more generally^7^. Nevertheless, a central question for mitigating the possible long-term effects of apnoea is knowing which events lead to significant cardiorespiratory instability. A common definition of apnoea is a pause in breathing lasting more than 20 seconds, or at least 10 seconds with associated bradycardia or desaturation^8^. Whilst there is a lack of a consistent clinical definition of apnoea, which leads to variations in clinical practice^9^, many neonatal units will use a set duration apnoea alarm threshold (such as 20 seconds) applied across all infants. But is a set duration alarm threshold appropriate for all infants? Are some infants more likely to experience cardiorespiratory instability from short apnoeas, whereas others may be relatively more resilient to prolonged pauses in breathing? Can we predict which infants will experience apnoea-related cardiorespiratory instability?

To fully address these questions and move towards personalised alarm limits, we must consider (1) which factors influence the risk of cardiorespiratory instability from apnoea (i.e., given an apnoea of any duration are some infants more likely to experience cardiorespiratory instability), (2) which factors modulate the relationship between cardiorespiratory instability and apnoea duration (i.e., is a different relationship between cardiorespiratory instability and apnoea duration observed in different infants), and (3) whether models which can predict risk of cardiorespiratory instability can be developed. Overall, the extent of cardiorespiratory instability, including change in heart rate, oxygen saturation, and cerebral oxygenation during apnoea are significantly correlated with the duration of apnoea^2,10–16^. However, considerable variability occurs in these relationships, with some short apnoeas resulting in substantial changes in cardiorespiratory stability and conversely some longer apnoeas not resulting in a change^2,17^. Type of apnoea (central, obstructive or mixed)^18–20^ and the use of theophylline^18^ (with limited evidence) have been shown to affect the relationship between apnoea duration and cardiorespiratory changes. Moreover, through meta-analysis, we recently identified postmenstrual age (PMA) as an important modulator: younger neonates were more likely to experience oxygen desaturation in response to relatively brief apnoea episodes^2^. Other factors that modulate the relationship are unknown. Gestation age at birth, haemoglobin concentration, type of respiratory support, caffeine treatment, and apnoeas that are clustered together may increase the overall risk of cardiorespiratory instability, but whether these factors also act as modulators has not been considered^21^. Identifying relevant modulators in the relationship between apnoea duration and cardiorespiratory instability will help determine which infants are most at risk of significant cardiorespiratory changes from apnoea, ultimately leading to the development of predictive models and from these personalised alarm limits.

The aim of this study was to (1) explore which factors either influence the risk of cardiorespiratory instability from apnoea and/or modulate the relationship between cardiorespiratory instability and apnoea duration, and (2) determine whether clinical, demographic and dynamic (i.e., changing physiological parameters which vary within an individual) factors can predict which apnoeas will result in cardiorespiratory instability (bradycardia and/or desaturation). To fully characterise the relationship between apnoea and cardiorespiratory instability, we used a broad approach, including apnoeas defined as pauses in breathing of at least 5 seconds.

## 2. Methods

### 2.1 Study design and eligibility criteria

Infants were selected from a database of data recorded by our research group between 2017 and 2025 at the John Radcliffe Hospital, Oxford University Hospitals NHS Foundation Trust, Oxford, UK. Ethical approval was obtained from the National Research Ethics Service (references: 12/SC/0447, 23/NW/0155, and 19/LO/1085), and informed written parental consent was obtained prior to each study. All studies were in accordance with the standards of the Declaration of Helsinki and adhered to Good Clinical Practice guidelines.

We included studies in which continuous vital-sign recordings were obtained from preterm infants. The eligibility criteria for recording sessions and apnoea episodes were:

- Age: Postmenstrual age (PMA) <37 weeks at the time of recording (study focus: preterm infants).
- Mode of ventilation: Infants were included if they were self-ventilating in air (SVIA) or receiving low-flow or high-flow oxygen therapy (the latter delivered via Vapotherm Precision Flow). Infants receiving other forms of respiratory support were excluded to ensure that extracted apnoea episodes and their physiological responses were not influenced by mechanical ventilation.
- Apnoea detection: A recording was included only if at least one apnoea (pause in breathing >5 seconds) was detected.
- Stable cardiorespiratory baseline: Apnoea episodes were excluded if, during the 90 seconds before apnoea onset, oxygen saturation fell below 85%, heart rate fell below 100 bpm, or heart rate exceeded 200 bpm. This requirement ensured that included apnoeas began from a reasonably stable cardiorespiratory baseline so subsequent changes could more plausibly be attributed to the apnoea itself.
- Medication: Infants receiving antibiotics, opioids, morphine or fentanyl at the time of the apnoea episode were excluded.

Of note, some infants receiving high-flow oxygen received this using automated oxygen control (delivered via the Vapotherm oxygen assist module (OAM)), which automatically titrates the FiO_2_ according to the infant’s oxygen saturations to achieve a preset target saturation. This was initiated in our unit from 2021 onwards and is locally recommended in all preterm infants <32 weeks (until 36 weeks PMA) and infants with PPHN (persistent pulmonary hypertension of the newborn), under clinician discretion. For the rest of the infants receiving high-flow oxygen, FiO_2_ is manually controlled by clinical staff.

### 2.2 Participants and recordings

Based on age, ventilation mode and medication criteria, there were 277 recordings from 156 infants eligible for inclusion in the study. After next including the apnoea criterion, the eligible sample size decreased to 257 recording sessions in 146 infants. 181,511 eligible apnoeas were detected from these included recordings (detailed apnoea detection rules are in Section 2.4.1).

The 257 included recording sessions totalled 23,854 hours of vital signs recordings (median: 50.2, range: 0.7–634.3 hours per recording). Of the included 146 infants, 99 had one recording session only, 19 had two sessions, and 28 had three or more. The length of recording varied considerably due to differences in study design at the time of data collection. Some infants took part in a research study where multiple test occasions were conducted, with vital signs recorded continuously throughout the hospital stay. This subset included 39 infants, contributing 133 test occasions, with a median recording length of 155.9 hours per session (range: 0.9–634.3 hours). Other research studies recorded single epochs of vital signs, with a median duration of 19.6 hours (range: 0.7–242.0 hours). Nine recordings from nine unique infants had a recording duration longer than two weeks. To ensure that these extended recordings did not disproportionately influence the results, we conducted a sensitivity analysis, repeating the statistical analysis using only those recordings with a duration of less than two weeks (Supplementary Table 1). No meaningful differences were observed between the findings from the complete dataset and those from the restricted dataset. Therefore, all recordings were included in the analysis regardless of recording length.

### 2.3 Data collection

Each infant had continuous vital signs monitoring as part of standard clinical care. Monitoring was conducted using Philips IntelliVue MX800 or MX750 monitors, and data were continuously downloaded using an electronic data capture system (iXtrend, ixitos, Germany). Vital signs including heart rate, oxygen saturation, and respiratory rate (as calculated by the monitor) were recorded at a sampling rate of 0.97 Hz. Additionally, high-resolution physiological signals were captured: the electrocardiograph (ECG), used to determine heart rate, was sampled at 250 Hz and recorded via three chest electrodes; the impedance pneumography (IP), used to assess respiration, was sampled at 62.5 Hz via the same chest electrodes; and the photoplethysmography (PPG), used to monitor oxygen saturation and pulse, was sampled at 125 Hz from a probe placed on the infant’s hand or foot.

Relevant demographic and clinical information for each infant was extracted from medical records for each recording session. The demographic and clinical indicators investigated as potential modulating factors are defined as follows:

- Sex: the biological sex at birth.
- PMA: postmenstrual age in weeks at the time of recording.
- GA: gestational age in weeks at the time of birth.
- Weight z-score: the infant’s weight z-score at the time of recording, relative to their PMA, calculated according to the guideline published by the RCPCH^22^, using the UK-WHO preterm weight data^23^ as reference.
- Ventilation: the mode of respiratory support the infant was receiving at the time of recording (self-ventilating, low flow, or high flow).
- Apgar (5 min): Apgar score at five minutes after birth^24^.

The dynamic features considered as modulators are defined as follows:

- HR baseline: the mean heart rate during the baseline period (90 seconds preceding the apnoea).
- SpO_2_ baseline: the mean oxygen saturation during the baseline period (90 seconds preceding the apnoea).
- Time since last apnoea: the interval between the end of the most recent apnoea and the start of the apnoea of interest in seconds.
- 5-min apnoea rate: the number of apnoeas within the last 5 minutes preceding an apnoea.
- Duration: the duration of the apnoea in seconds.

### 2.4 Data analysis

#### 2.4.1 Identification and classification of apnoea events

Pre-processing and analyses were performed in MATLAB (ver. 2024a; MathWorks Inc., Natick, USA) and Python (ver. 3.11.5; Python Software Foundation, Wilmington, USA). Inter-breath intervals (IBIs) were identified from the impedance pneumograph (IP) signals, by applying a previously validated algorithm^25^. Briefly, the algorithm first filters the signal to reduce the noise introduced by movements and cardiac activity. It then identifies individual breaths using an adaptive amplitude threshold. Finally, the algorithm eliminates false positive apnoeas (e.g. due to shallow breathing or poor signal) using a Support Vector Machine (SVM) classifier (trained on a data set independent from the current data). The machine learning classifier developed in this algorithm was validated on IBIs greater than 15 seconds only^25,26^. Since we were also interested in short pauses in breathing in this study, IBIs between 5 and 15 seconds were filtered using a new independent machine learning algorithm (Supplementary Methods A).

All IBIs lasting between 5 and 40 seconds that were identified by the machine learning algorithms as true apnoeas were included in this study as apnoea events. IBIs exceeding 60 seconds were manually checked before including as apnoea events, as clinical experience indicates these are more likely to be false positives than true apnoea episodes. IBIs between 40 and 60 seconds were manually reviewed if they were associated with less than 20% drop in either heart rate or oxygen saturation, as these were also more likely to be false positives. During the manual review, both the original and filtered IP signals were examined to determine whether the detected pauses in breathing were due to potential connection issues causing abnormal signals, or genuine apnoeas characterised by a clear cessation of breathing.

#### 2.4.2 Definition of cardiorespiratory outcomes and analysis windows

Desaturation events were defined as a decrease in oxygen saturation to below 85% lasting for at least five consecutive seconds. Bradycardia was defined as a decrease in heart rate of more than 30% from baseline lasting for at least five consecutive seconds. Whilst bradycardia in neonates is often defined as a heart rate below 100 or 110 beats per minute, older infants have baseline heart rates close to this range without any physiological compromise. Consequently, using a definition of bradycardia as less than 100 beats per minute meant that bradycardia rates increased with increasing PMA, distorting results (Supplementary Methods B, Supplementary Figure 1). In contrast, when using a definition of 30% from the baseline, consistent with definitions used in other literature^27,28^, rate of bradycardia does not change with PMA (Supplementary Figure 1).

The baseline period was defined as the 90 seconds preceding the start of each apnoea episode, and the post-event period was defined as the interval from the onset of an apnoea episode to 10 seconds after its end. These time frames were chosen to capture delayed cardiorespiratory effects, as changes in heart rate and oxygen saturation may lag behind apnoea due to physiological response times and pulse oximeter delay, and are typically fully captured within 10 seconds after the end of the apnoea^16^. The window was not extended further to minimise potential confounding and to ensure that observed changes were associated with the apnoea.

#### 2.4.3 Resampling and capping

As the number of apnoeas per infant and per recording had a very wide range, to ensure the results were not overly influenced by infants with higher numbers of apnoeas, we generated 500 resampled datasets by randomly sampling apnoea events within each recording, capping the number at 200 per recording. This approach reduced the influence of recordings with large numbers of events while allowing estimation of parameter variability. All the statistical and machine learning analysis were performed on resampled datasets and aggregated using the methods below. A flowchart of the analysis steps and modelling procedure is presented in Supplementary Figure 2.

#### 2.4.4 Descriptive analysis of physiological responses by apnoea duration

For visualisation purposes, the associated physiological responses were compared with apnoea duration using sliding windows of 5 seconds’ duration with 70% overlap. Windows with fewer than 20 data points, or derived from fewer than three unique recordings, were excluded from the plots. Within each window, summary statistics were estimated on 500 resampled datasets. For each resampled dataset and duration window, the mean change in heart rate, mean minimum oxygen saturation, and the proportion of apnoeas associated with bradycardia and/or desaturation (apnoea with bradycardia/desaturation) were calculated. Across replicates, the distribution of each statistic was used to derive the mean estimate and corresponding 95% confidence intervals. In addition, Pearson’s correlation coefficients were computed to quantify the linear association between apnoea duration and each physiological outcome.

#### 2.4.5 Mixed-effects modelling of factors associated with apnoea-related instability

This analysis aimed to distinguish between overall effects of each factor on the risk of cardiorespiratory instability (i.e., the likelihood of an apnoea of any duration being associated with bradycardia and/or desaturation), and interaction effects indicating whether a factor modulates the relationship between apnoea duration and instability. In other words, we examined both general risk differences between infants and whether specific characteristics alter the duration–response association.

Logistic mixed-effects models were employed on the resampled datasets to analyse the influence of factors on the relationship between apnoea duration and cardiorespiratory responses. The two-way interaction terms between apnoea duration and each factor of interest (sex, PMA, GA, weight z-score, ventilation, Apgar score at 5 min, heart rate baseline, oxygen saturation baseline, time since last apnoea, and the number of apnoeas in the past 5 minutes) were included as fixed effects in separate models. Recording session ID and infant ID were included as random effects to account for repeated measures within infants. The response variable was binary, defined as 0 for apnoea episodes not associated with bradycardia or desaturation, and 1 for apnoea with bradycardia and/or desaturation. The model coefficients and associated p-values from all replicates were extracted and combined across resampled datasets. For each fixed-effect term, we calculated the mean coefficient (representing the average effect size). The mean coefficient corresponded to the log-odds ratio, and its exponential transformation was used to obtain the odds ratio. We also derived the 95% confidence intervals (CIs) for each coefficient and determined whether the aggregated coefficients were significant by assessing whether the 95% CI crossed the null value. In addition, figures were generated using the resampled datasets in a similar manner to that described in Section 2.4.5, but stratified by each feature. Stratification thresholds were selected to reflect either clinically meaningful cut-offs or values close to the median of the respective feature.

#### 2.4.6 Machine-learning prediction of apnoea-related instability

We constructed binary classification models using the eXtreme Gradient Boosting (XGBoost) algorithm to predict apnoea-related bradycardia and/or desaturation events using clinical/demographic and dynamic features. Model development was restricted to a complete-case subset with no missing feature values (N = 153,947 apnoeas), and apnoea episodes with missing features were excluded (N = 27,564). The response variable was binary, again defined as 0 for apnoea episodes not associated with bradycardia or desaturation (N = 146,053; 94.9%) and 1 for apnoea associated with bradycardia and/or desaturation (N = 7,894; 5.1%). The dataset was partitioned at the subject level into a training set (N = 131,638) and a held-out test set (N = 22,309), stratified by apnoea duration, postmenstrual age (PMA), and the response variable. All recording sessions from the same infant were allocated in the same set. Sub-models were trained on 10 independently resampled versions of the training data and combined via averaging. Model training involved tuning of hyperparameters (Supplementary Method C) via random search with 500 iterations, using internal 10-fold cross-validation on the training set. Out-of-fold (OOF) predicted probabilities were generated on the training set. Classification thresholds were selected by evaluating all candidate thresholds on the OOF predictions and choosing the threshold that maximised balanced accuracy. An ensemble model was constructed from the models trained on all resampled versions of the training data. Model performance was then evaluated on the independent test set using balanced accuracy and area under the receiver operating characteristic curve (AUROC).

#### 2.4.7 Permutation testing of model performance

To evaluate whether the XGBoost classification model performs significantly better than chance, permutation tests were conducted for every model. This method involves generating a null distribution of predictive performance by training the classifier on data (1,000 permutations) in which the class labels in the training set were randomly permuted. For each permutation, a new XGBoost model was trained and evaluated on the same test set. The p-value was then calculated as the proportion of permuted models that achieved an accuracy equal to or greater than that of the original model. The evaluation was done on the fixed, held-out test set, which was not used in any stage of model training or tuning.

#### 2.4.8 Model-interpretation analysis of predicted risk profiles using synthetic data

To characterise the combined feature profiles associated with higher and lower predicted risk of apnoea-related bradycardia/desaturation we used the reduced demographic/clinical model to identify the types of infants classified as relatively high or low risk within each apnoea-duration band. This analysis served as a model-interpretation step, aiming to clarify the clinical meaning of the multivariable predictions. It is profile-based rather than variable-based and therefore complements, rather than replaces, the mixed-effects analyses of individual factors and interactions. Moreover, the purpose was descriptive and interpretative, not causal, and should not be considered a new independent test of the contribution of individual variables.

Synthetic data were generated by randomly sampling each feature from a uniform distribution 10,000 times. Ventilation mode was sampled categorically, with the probability of each mode set to match the observed distribution of ventilation modes among infants of the corresponding age and sex in the real dataset. The optimised model which only used demographic and clinical variables was applied to the synthetic dataset to generate predicted risk scores for apnoea with bradycardia/desaturation for each observation. Predicted events were stratified by apnoea duration into three categories: 5–10 seconds, 10–20 seconds, and ≥20 seconds. Within each duration category, comparisons were conducted between high- and low-risk groups, defined as the highest and lowest 10% of predicted risk, respectively. To avoid pseudo-replication arising from multiple observations per recording, all risk-group comparisons were performed at the subject level. Specifically, subjects were ranked according to their mean predicted risk within each duration category, and those in the highest and lowest 10 percentiles were compared. The comparisons were done on demographic or clinical variables only. For continuous variables, group differences were assessed using Welch’s two-sample t-test, while categorical variables were compared using Fisher’s exact test or Pearson’s χ² test, as appropriate. All statistical tests were two-sided, with significance evaluated using conventional thresholds (p < 0.05, p < 0.01, p < 0.001).

## 3 Results

### 3.1 Cardiorespiratory instability relates to apnoea duration but with wide variation

A total of 181,511 apnoeas (pauses in breathing for at least 5 seconds) were included in the analysis, from 257 recording sessions in 146 infants (Figure 1). An average of 287 apnoeas (median, interquartile range: 40–914) occurred per recording, and the average number of apnoeas per hour was 6.8 (median, interquartile range: 3.7–13.0). Demographic and clinical information on the included infants are provided in Table 1, and the characteristics of all apnoea episodes are given in Table 2.

**Figure 1:**
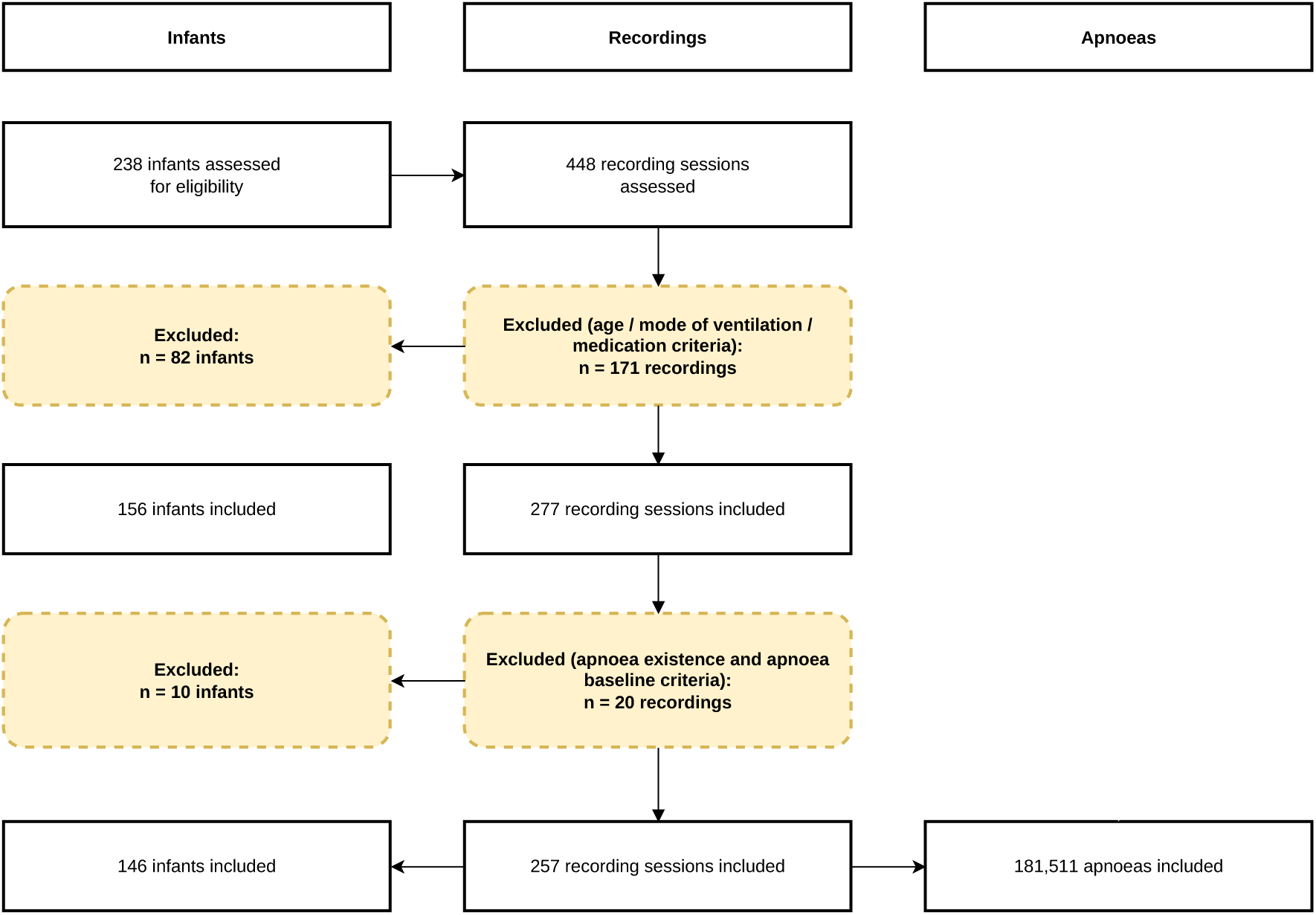
Data collection flow chart.

**Table 1:** Demographic and clinical variables at the start of all test occasions. Values expressed as median (range) or percentage. Variables that do not change over time are reported per infant, whereas time-varying variables are reported per recording session. SVIA: self-ventilating in air.

| Variables | Per infant (N = 146) |
| --- | --- |
| Gestational age at birth (weeks) | 28.9 (23.0 – 35.0) |
| Sex - Female | 44.5% |
| Apgar score 5 min after birth | 9 (3 – 10) |
| Variables | Per recording session (N = 257) |
| Recording length (hours) | 50.2 (0.7 – 634.3) |
| Postmenstrual age at study (weeks) | 33.2 (26.3 – 36.9) |
| Weight z-score at study | -0.8 (-3.7 – 2.3) |
| Number of apnoeas per hour | 6.8 (0.0 – 57.5) |
| Ventilation - SVIA | 45.1% |
| Ventilation - Low Flow | 14.4% |
| Ventilation - High Flow | 40.5% |

**Table 2:** Details of all apnoea episodes. Values expressed as median (range). Bpm: beats per minute.

| Variables | All apnoeas (N = 181,511) |
| --- | --- |
| Apnoea duration (seconds) | 6.8 (5.0 – 117.3) |
| Heart rate baseline (bpm) | 158.2 (100.0 – 200.0) |
| SpO<sub>2</sub> baseline (%) | 96.3 (85.0 – 100.0) |
| Number of apnoeas in last 5 minutes | 2 (0 – 24) |
| Time since last apnoea (seconds) | 84.9 (2.0 – 954614.2) |
| Minimum heart rate post-event (bpm) | 150.0 (29.0 – 209.0) |
| Minimum oxygen saturation post-event (%) | 94.2 (1.9 – 100.0) |
| Change in heart rate post-event (%) | -4.3 (-82.4 – 60.0) |
| Change in oxygen saturation post-event (%) | -1.6 (-98.0 – 17.5) |

Overall, heart rate and oxygen saturation significantly decreased with increasing apnoea duration (heart rate: r = −0.84, p < 0.0001; oxygen saturation: r = −0.96, p < 0.0001; Figure 2A-B). Similarly, the proportion of apnoeas with bradycardia/desaturation significantly increased with apnoea duration (r = 0.82, p < 0.0001, Figure 2C). Nevertheless, there was wide variation in cardiorespiratory changes for apnoeas of a given duration; even with short pauses in breathing of 5-10 seconds, bradycardia/desaturation occurred in 3.6% (a total of 5617 short pauses in breathing from 95 infants, on average 6.34 per day in these infants). Conversely, for 61.2% of apnoeas (N = 2214) longer than or equal to 20 seconds no bradycardia or desaturation occurred.

**Figure 2:**
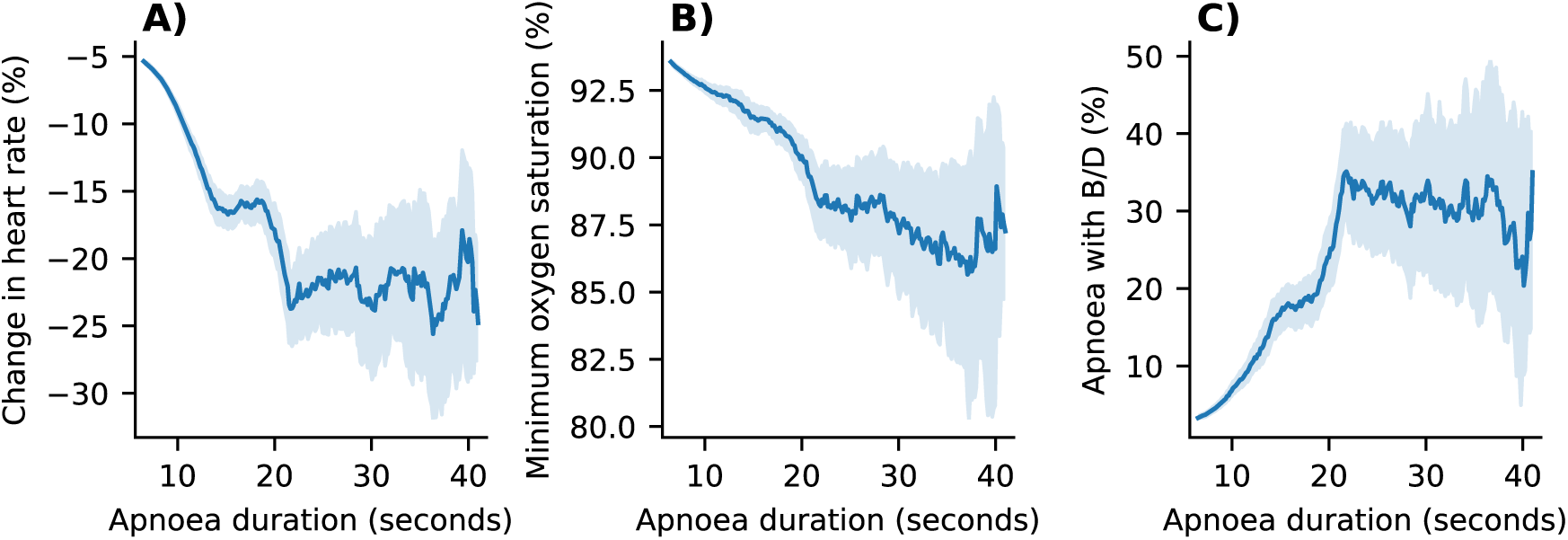
Physiological responses compared with apnoea duration. (A) Change in heart rate (%) relative to baseline compared with apnoea duration. (B) Minimum oxygen saturation (%) observed during apnoeas compared with apnoea duration. (C) Proportion (%) of apnoeas associated with bradycardia and/or desaturation (B/D) compared with apnoea duration. Apnoeas were grouped by duration using overlapping windows (70% overlap). Solid lines represent the mean estimates across resampled datasets, and shaded regions indicate the corresponding 95% confidence intervals.

### 3.2 Demographic and clinical factors increase risk of cardiorespiratory instability

PMA at study, GA at birth, mode of ventilation, weight z-score and sex all had a significant effect overall (i.e., irrespective of apnoea duration) on the likelihood of apnoea-related bradycardia and/or desaturation (linear mixed effects models, Table 3, Figure 3 panels A and C). Infants with a younger PMA, younger GA, receiving high flow therapy at the time of study, smaller babies and those who are male were more likely to have substantial cardiorespiratory instability (Figure 3, columns A and C). Apgar score at birth did not have a significant impact on the likelihood of bradycardia/desaturation (Table 3, Figure 3).

**Figure 3:**
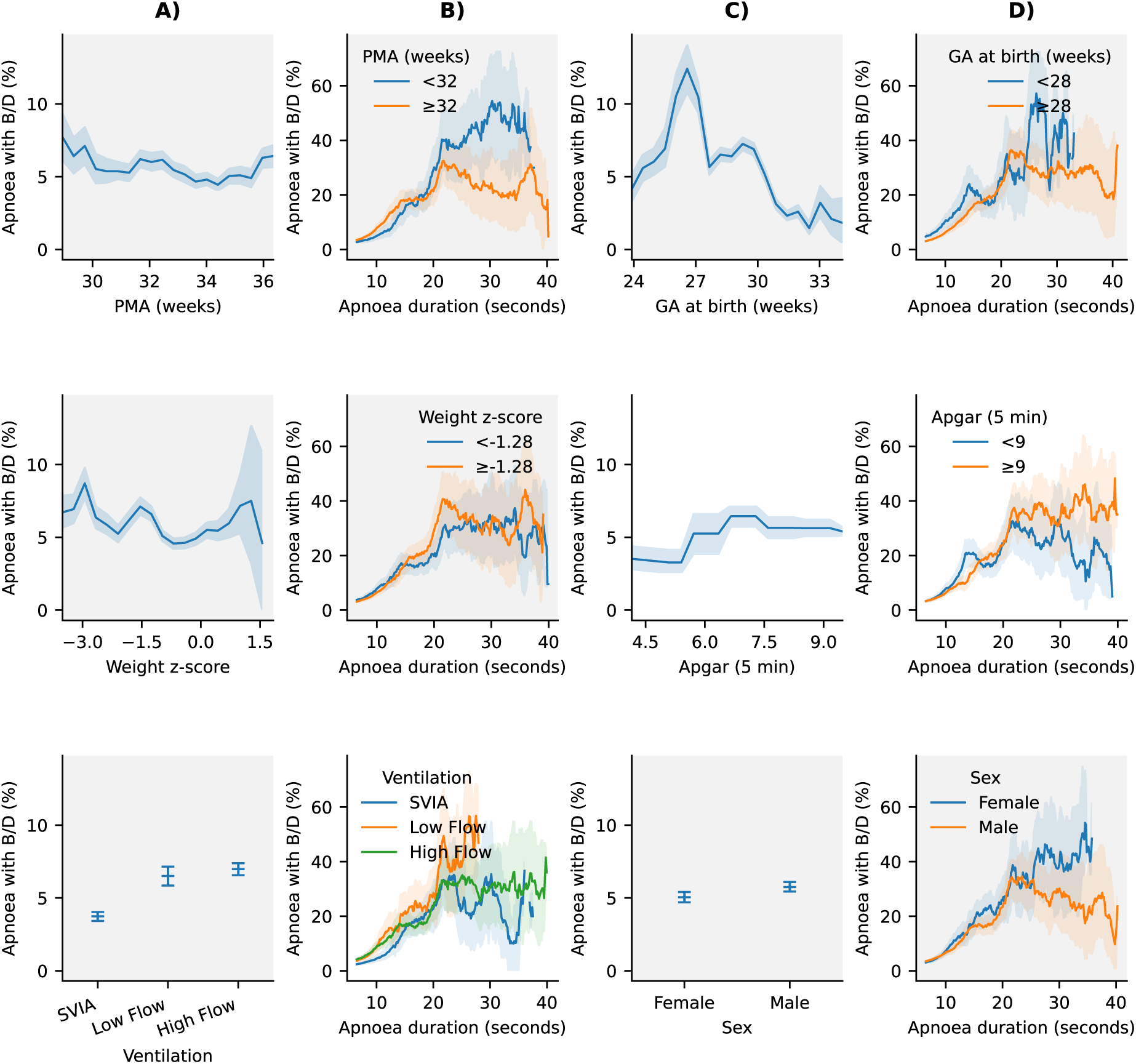
Influence of demographic and clinical factors on apnoea-related bradycardia and/or desaturations (B/D). Columns A) and C) illustrate the proportion of apnoeas associated with bradycardia/desaturation compared with demographic and clinical variables. Columns B) and D) show the relationship between apnoea duration and the proportion of apnoeas with bradycardia/desaturation, stratified by the same variables (indicating whether the factor modulates the relationship between apnoea duration and occurrence of bradycardia/desaturation, with stratification thresholds selected to reflect either clinically meaningful cut-offs or values close to the median of the respective feature). Apnoeas were grouped by duration using overlapping windows. Solid lines represent mean estimates across resampled datasets, and shaded regions indicate the corresponding 95% confidence intervals. Background grey shading indicates significant factors (see Table 3).

**Table 3:**
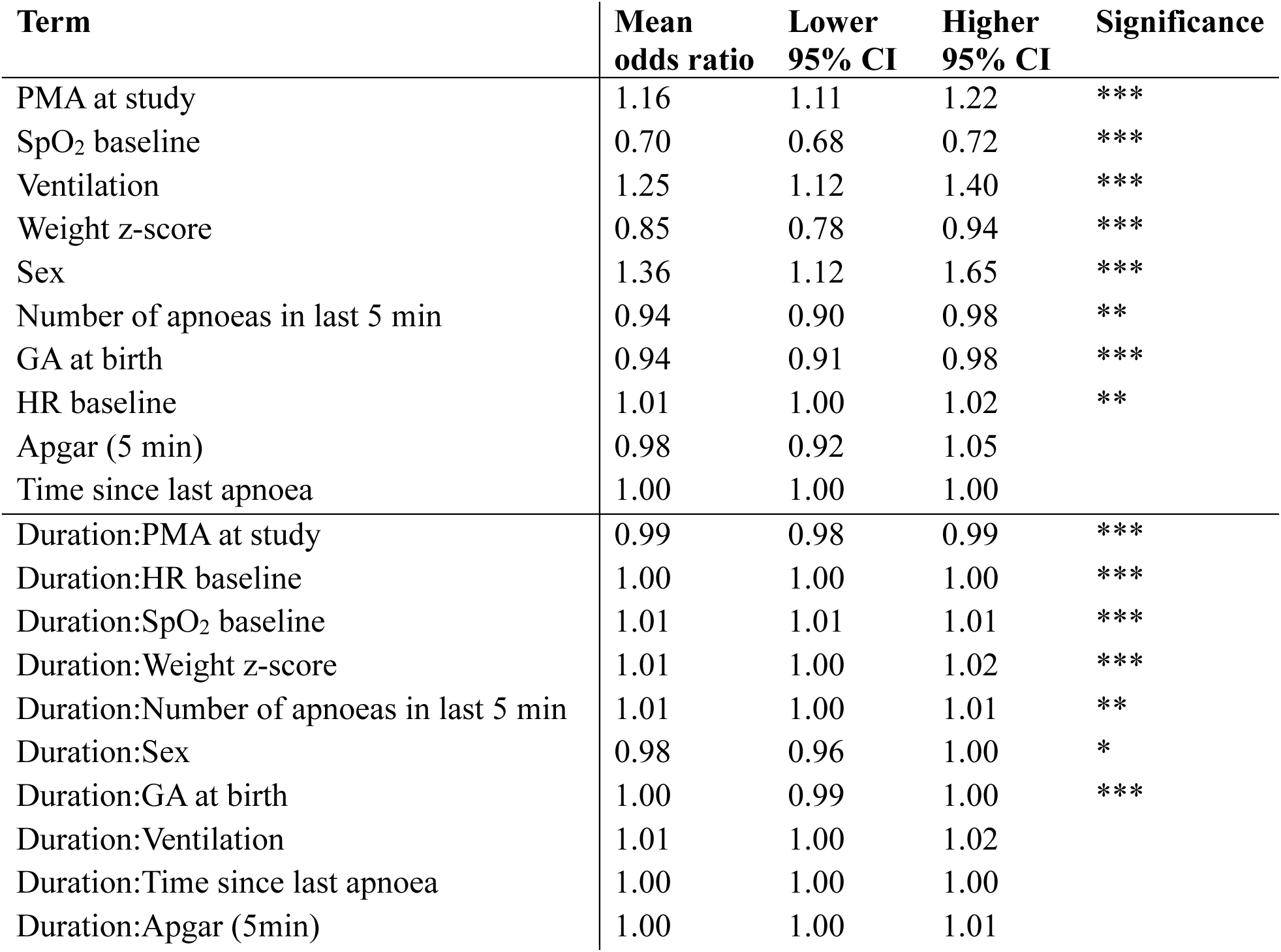
Results from univariate mixed-effects models examining the effects of individual factors and their interactions with apnoea duration on the likelihood that an apnoea is associated with bradycardia and/or desaturation. PMA = postmenstrual age, HR = heart rate, GA = gestational age, CI = confidence interval. * significant at the 95 % CI, ** significant at the 99 % CI, *** significant at the 99.9 % CI.

We also considered which factors significantly modulated the relationship between apnoea duration and the proportion of apnoeas with substantial cardiorespiratory instability. PMA at study, weight z-score, sex, and GA at birth were all significant modulating factors (Table 3, Figure 3 panels B and D). In contrast, mode of ventilation and Apgar score were not significant modulating factors.

### 3.3 Multiple dynamic features increase the risk of cardiorespiratory instability

Baseline heart rate, baseline oxygen saturation, and the number of apnoeas in the preceding five minutes all had a significant effect on the likelihood of apnoea-related bradycardia/desaturation (linear mixed-effects models; Table 3, Figure 4 panels A and C). Infants with lower baseline oxygen saturation values and a greater number of apnoea episodes within the preceding five minutes were more likely to experience substantial cardiorespiratory instability (Figure 4, panels A and C). These variables were also significant modulators of the relationship between apnoea duration and the proportion of apnoeas associated with bradycardia/desaturation (Table 3; Figure 4, panels B and D). In contrast, time since the last apnoea did not significantly affect the likelihood of bradycardia/desaturation and was not a significant modulating factor. This may be because the relationship between apnoea history and physiological responses is not linear and can be influenced by multiple other factors, such as the duration and severity of the preceding apnoea.

**Figure 4:**
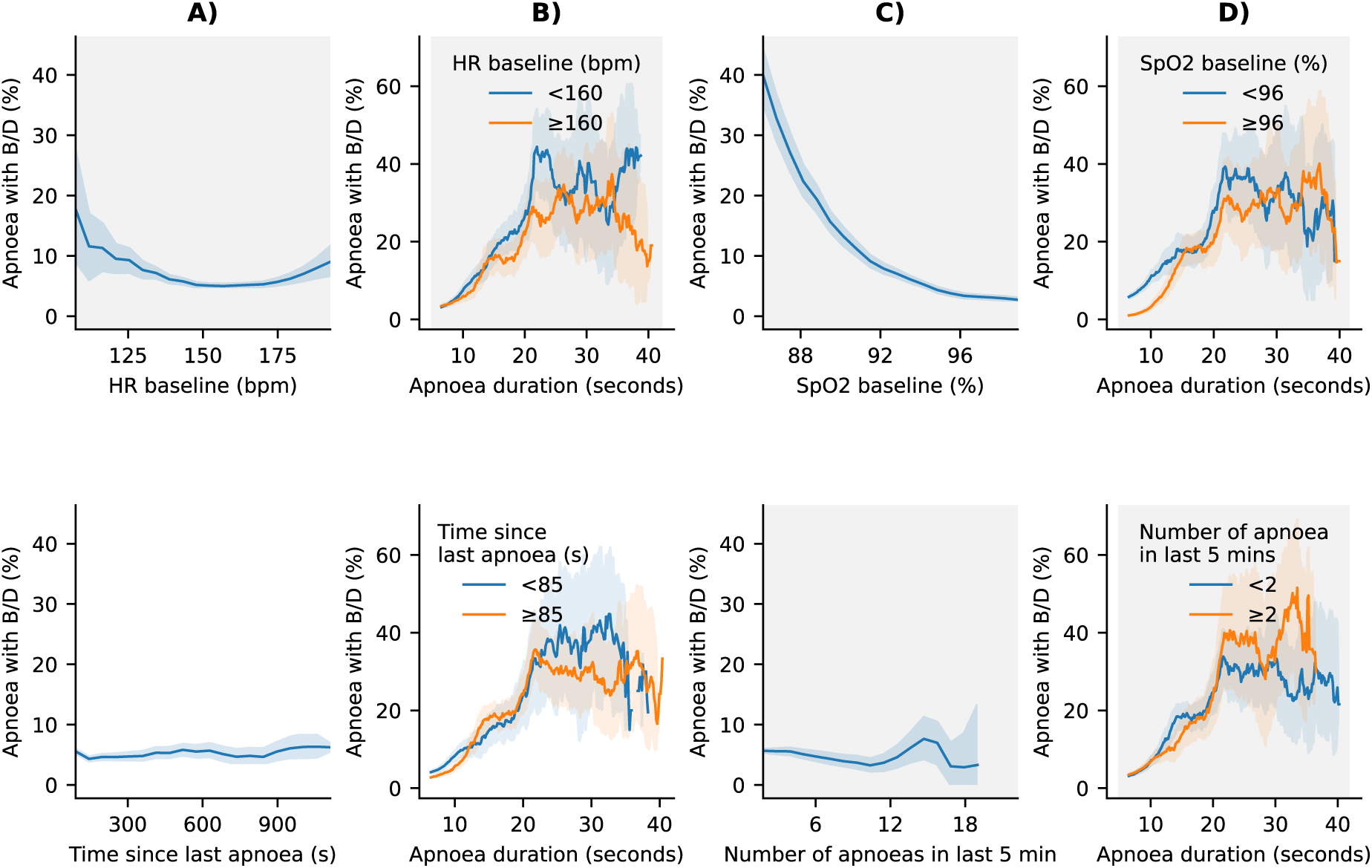
Influence of dynamic variables on apnoea-related bradycardia and/or desaturations (B/D). Columns A) and C) illustrate the proportion of apnoeas associated with bradycardia/desaturation compared with key dynamic variables. Columns B) and D) show the relationship between apnoea duration and the proportion of apnoeas with bradycardia/desaturation, stratified by the same variables (indicating whether the factor modulates the relationship between apnoea duration and occurrence of bradycardia/desaturation, with stratification thresholds selected to reflect either clinically meaningful cut-offs or values close to the median of the respective feature). Apnoeas were grouped by duration using overlapping windows. Solid lines represent mean estimates across resampled datasets, and shaded regions indicate the corresponding 95% confidence intervals. Background grey shading indicates significant factors (see Table 3).

### 3.4 Cardiorespiratory instability from apnoea can be predicted using machine learning

As a first step towards personalised alarm limits, we investigated if it is possible to predict apnoeas associated with bradycardia/desaturation. We included all demographic, clinical and dynamic factors as inputs to the model. Following random search optimisation, the best-performing model achieved balanced OOF train accuracy of 75.6% and a balanced test accuracy of 75.8% (Figure 5A), which was significantly greater than chance (permutation test, p < 0.0001). The model correctly identified 75.2% of apnoea events associated with bradycardia/desaturation, and 76.4% of apnoeas not associated with bradycardia/desaturation (Figure 5A). The AUROC was 0.83 on the held-out test set.

**Figure 5:**
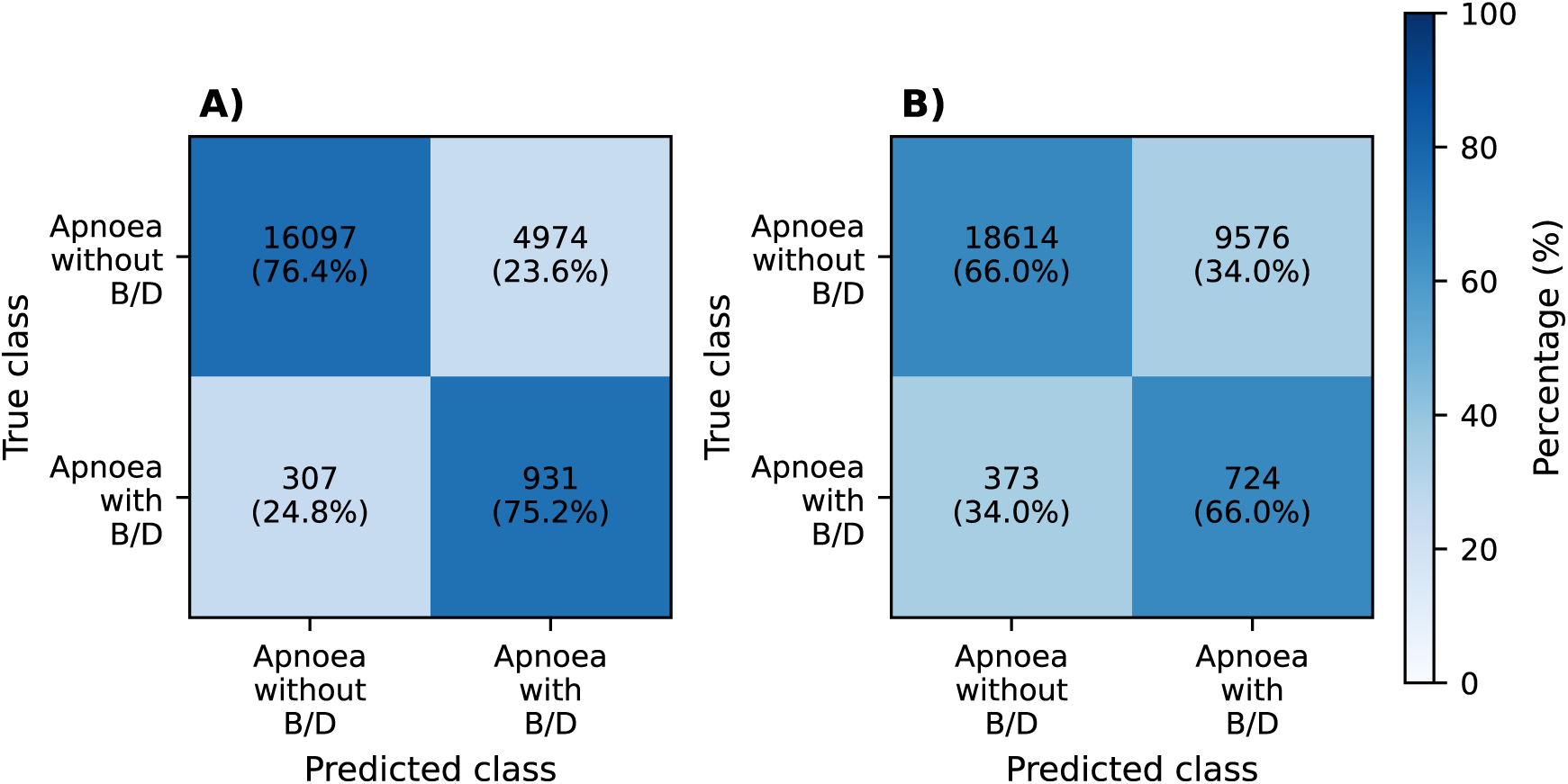
Confusion matrices of the XGBoost model prediction outcomes. Panel A shows results from the optimised model trained using all features, whereas panel B shows results from the optimised model trained using demographic and clinical features only (i.e., when not including the dynamic features). Numbers in each cell indicate the count of apnoea events in each category. B/D denotes bradycardia and/or desaturation.

Clinically, it is not always possible to obtain dynamic features, and adjusting apnoea alarm thresholds based on changing physiological information can be impractical. Therefore, we also investigated the possibility of using only demographic and clinical variables included as features, as these are relatively stable within an infant. A separately trained model achieved a balanced OOF train accuracy of 62.8% and a balanced test accuracy of 66.0% (Figure 5B), which was still significantly better than chance (permutation test, p < 0.0001). The AUROC was 0.72 on the held-out test set. To identify the profiles associated with high and low risk for cardiorespiratory instability during apnoea, this second model was used to predict the risk of having apnoea-related bradycardia/desaturation across a synthetic dataset. Synthetic profiles in the highest and lowest 10% of predicted risk for apnoea associated with bradycardia and/or desaturation at durations between 5 and 10 seconds differed significantly in GA at birth, PMA at the time of the study, sex, Apgar score 5 minutes after the birth, and ventilation mode (Table 4). Males, lower gestational age, lower postmenstrual age, lower Apgar scores, or receiving high-flow respiratory support are associated with a higher risk of experiencing short apnoea episodes associated with bradycardia and/or desaturation. Pairwise heatmaps of these significant features were generated to illustrate how predicted risk varies across these variables (Figure 6). Feature comparisons between high- and low-risk groups for longer apnoea durations are presented in Supplementary Tables 2-3 and Supplementary Figures 3-4.

**Figure 6:**
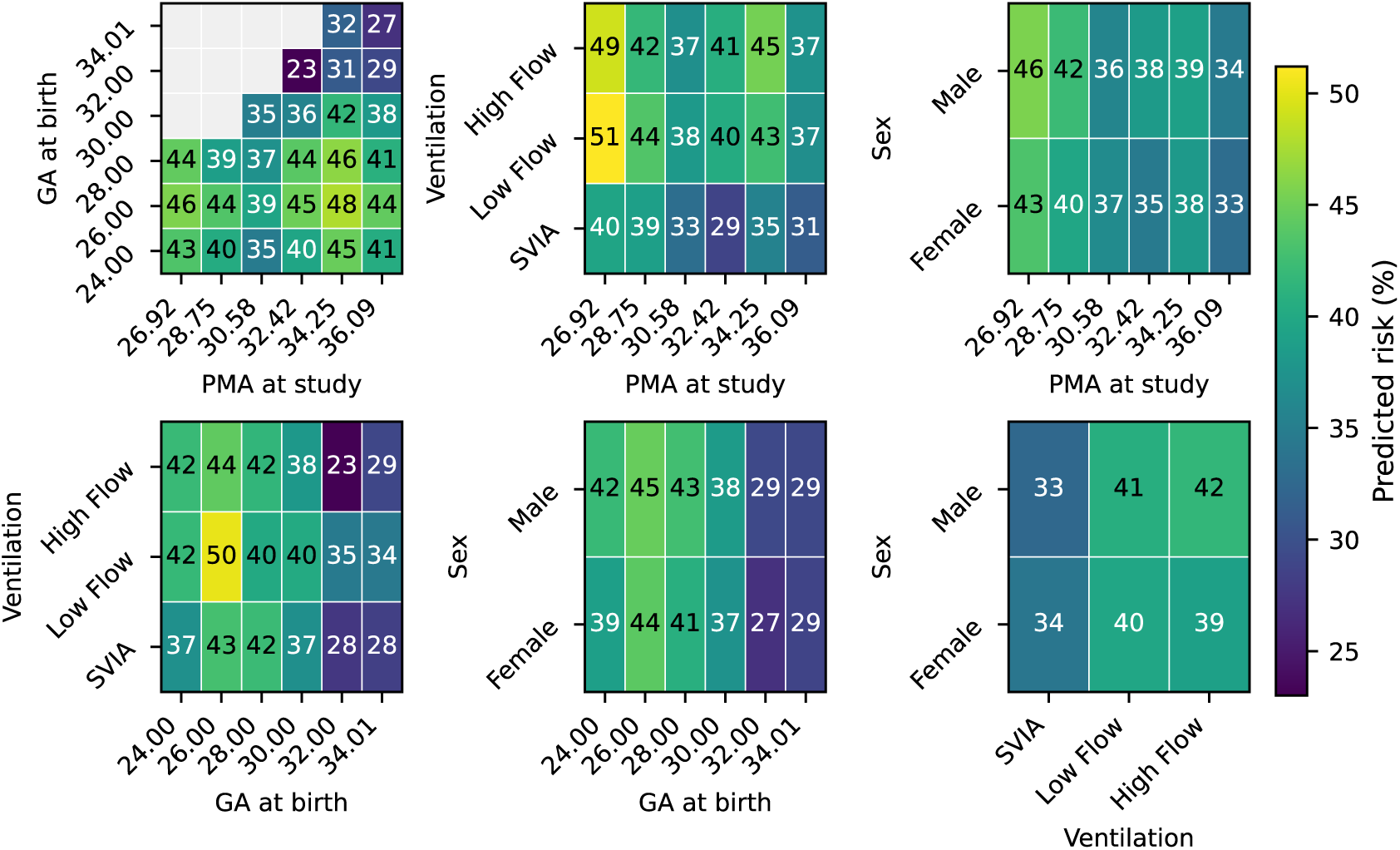
Pairwise heatmaps of the predicted risk of apnoea-related bradycardia and/or desaturation in synthetic data. Risk was predicted for all apnoeas with durations between 5 and 10 seconds using the optimised XGBoost model. Values shown in each heatmap cell (colour-coded and numerically annotated) represent the mean predicted risk for the corresponding feature combinations.

**Table 4:** Comparison of model features between apnoeas with the highest and lowest 10% predicted risk of bradycardia and/or desaturation in synthetic data. Features were compared between high- and low-risk groups based on model-predicted risk for all apnoeas with duration between 5 and 10 seconds. PMA = postmenstrual age, GA = gestational age. Statistically significant differences are indicated as follows: * p < 0.05, ** p < 0.01, *** p < 0.001.

| Term | High-risk mean | Low-risk mean | p value | Significance |
| --- | --- | --- | --- | --- |
| PMA at study | 32.81 | 34.39 | $< 0.0001$ | *** |
| GA at birth | 27.59 | 31.72 | $< 0.0001$ | *** |
| Weight z-score | -0.61 | -0.66 | 0.65 |  |
| Apgar (5 min) | 3.11 | 6.19 | $< 0.0001$ | *** |
| Ventilation=SVIA | 0.35 | 0.75 | $< 0.0001$ | *** |
| Ventilation=Low Flow | 0.10 | 0.04 | 0.00037 | *** |
| Ventilation=High Flow | 0.55 | 0.21 | $< 0.0001$ | *** |
| Sex=female | 0.45 | 0.57 | 0.0010 | *** |
| Sex=male | 0.55 | 0.43 | 0.0010 | *** |

## 4 Discussion

This study investigated factors underlying cardiorespiratory instability from apnoea in preterm infants aged 26-36 weeks PMA. We identified multiple demographic, clinical and dynamic factors which influence the risk of bradycardia and/or desaturation. Moreover, apnoeas with bradycardia and/or desaturation could be predicted using machine learning.

In current clinical practice, apnoea alarms are commonly defined using uniform thresholds across all infants. Bedside monitor alarms typically trigger only when apnoea lasts longer than 20 seconds, and breathing pauses shorter than 10 seconds are often not classified as apnoea in either research or clinical practice. However, our findings suggest that a subset of infants may experience severe cardiorespiratory instability following even relatively brief respiratory pauses. The instability reaches a plateau for apnoeas longer than approximately 20 seconds (Figure 2), which may be because clinical interventions prevent further declines in heart rate and/or oxygen saturation. On the other hand, 60% of apnoeas longer than 20 seconds are not associated with bradycardia or desaturation. These findings highlight the potential value of a personalised definition of apnoea incorporating demographic characteristics and dynamic factors, such as recent apnoea history. With further validation, this framework could form the basis of a web-based clinical tool to support clinical decision making and individualised apnoea alarm thresholds. This could enable earlier detection of clinically significant apnoeas in more vulnerable infants, which could trigger earlier clinical assessment and management, and potentially help to mitigate the risk of long-term impacts of apnoea-associated hypoxaemia^6^. Understanding the risks of cardiorespiratory instability from apnoea in different infants could also be used when planning procedures where infants may be more likely to experience apnoea^29^. Whilst the model presented here could not be directly used in clinical practice to define a personalised apnoea alarm (due to limitations such as sample size and lack of external validation), our study nevertheless provides the foundations for such a tool.

We previously conducted a systematic review and meta-analysis of the relationship between apnoea duration, cardiorespiratory and cerebrovascular responses in preterm infants^2^. Consistent with this review, we found that cardiorespiratory instability was significantly correlated with apnoea duration but there is nevertheless wide variation in response. Little research to date has explored factors modulating the relationship between apnoea duration and cardiorespiratory instability^2^. Moreover, our systematic review was limited as many studies were conducted before 2000^2,30^, where treatment regimens differed, such as changing trends in the mode of respiratory support used^31^. The current study provides a rigorous assessment of modulating factors in a contemporary cohort, shedding light on why some infants are more at risk of cardiorespiratory instability from apnoea and conversely why some infants may be more resilient. Both gestational age (GA) at birth and PMA at study significantly affected the risk of cardiorespiratory instability, with younger infants at higher risk. The respiratory system and the body’s oxygen carrying capacity matures with increasing PMA. With advancing PMA, functional residual capacity increases, surfactant production is enhanced, respiratory muscles become stronger and alveoli development is more mature^32^. Moreover, the shift from predominantly foetal haemoglobin to adult haemoglobin may result in increased oxygen delivery to tissues^33^. These changes likely means that infants with older PMA are more resilient to longer duration apnoeas.

The age at which apnoeas stop is related to GA at birth, with infants born at younger GA experiencing apnoea until older PMA^7^. Our data show that when infants of younger GA at birth experience apnoeas, they are more likely to be associated with bradycardia and desaturation, and that GA at birth also modulates the relationship between apnoea duration and cardiorespiratory instability. This could in part be related to relative immaturity of lung function, which is known to persist after the initial neonatal period and could impact on cardiorespiratory response to apnoea, compared to their higher GA at birth counterparts^34^. Infants born at lower GA are also at increased risk of numerous pathologies, including anaemia of prematurity, intraventricular haemorrhage, infections, metabolic disturbances and temperature instability, all of which can both be pathological causes of apnoea but could also reduce infants’ ability to tolerate apnoeas^35–37^. Moreover, infants born at lower GA are likely to require prolonged mechanical respiratory support and may subsequently develop bronchopulmonary dysplasia (BPD), which can impair pulmonary gas exchange and could increase extremely preterm infants’ vulnerability to apnoea-associated desaturation^38,39^.

Our data show that infants who experience clusters of apnoeas are more vulnerable to cardiorespiratory instability with a subsequent apnoea. Clinically, short recurrent apnoeas can be seen in periodic breathing, defined as at least 3 pauses in breathing (lasting ≥ 3 seconds each), separated by less than 20 seconds each^40^. They are relatively common, particularly in preterm infants, and are generally considered benign and self-resolving^41^. However, previous studies have shown that clusters of brief apnoeas as short as 3 seconds may cause significant bradycardia and desaturation, and even changes in cerebral oxygenation and blood volume^21,42–45^. Our data add to this work and suggests that having multiple (even brief) apnoeas within a 5-minute period should alert clinicians to the increased chance of a subsequent desaturation/bradycardia. Previous studies have also found associations between multiple brief apnoeas (e.g. in sequential apnoeas and periodic breathing) and poorer neurodevelopmental outcomes at 6 months and 2 years of life, reinforcing the potential clinical seriousness of these seemingly insignificant events^42,46^.

Our study found an association between mode of respiratory support (SVIA, low flow and high flow oxygen) and the likelihood of apnoea-associated bradycardia/desaturation. The mode of respiratory support an infant receives may be influenced by a number of factors, including the infant’s overall clinical status and the presence of respiratory or other comorbidities (e.g. respiratory distress syndrome (RDS), BPD, pneumonia), some of which may also impair an infant’s ability to tolerate apnoeas without cardiorespiratory compromise. The choice of respiratory support is often driven by local clinical guidelines as well as clinician judgement, and as such this result needs to be considered cautiously in interpretation across different neonatal units. Moreover, this study also has several other limitations. The dataset included only central and mixed apnoeas and did not capture obstructive apnoeas as nasal air flow is not routinely monitored on our neonatal unit. Consequently, analyses stratified by apnoea type were not possible. Moreover, the majority of infants were older than 28 weeks PMA at the time of recording; evaluating cardiorespiratory instability risk in the youngest infants should be considered in a separate study. Conversely, as our hospital is a regional centre, preterm infants who are clinically stable are more likely to be discharged or transferred to other hospitals at an earlier postmenstrual age. As a result, infants with recordings available at late preterm ages in this dataset are more likely to represent a clinically unstable or complex subgroup, introducing potential bias into analyses involving PMA.

Finally, the set of potential modulating factors within this data set is not exhaustive. For example, we initially planned to investigate the impact of infection, however, the majority of our data was not recorded in infants with current infection and so we excluded these infants from analysis. The impact of anaemia, steroids, surfactant, other medication, concurrent respiratory pathologies (e.g. BPD, RDS) and cardiac pathologies (e.g. PDA) would also be important to consider and should be addressed in future predictive models to obtain a tool suitable for all infants. Despite these limitations, this study provides strong evidence that infant demographic characteristics, recent apnoea history, and baseline physiological state significantly influence the cardiorespiratory response to apnoea. The severity of apnoea-related instability varies considerably between individuals, and for some infants, respiratory pauses shorter than 10 seconds may already be related to a bradycardia or desaturation, suggesting that the definition of apnoea may warrant further consideration.

In summary, we demonstrate that multiple factors modulate the risk of bradycardia and/or desaturation from apnoea. Infants born at younger gestational ages, current younger postmenstrual age, and receiving high flow therapy were at higher risk than their older counterparts who were self-ventilating or receiving low flow therapy. Moreover, apnoea with bradycardia and/or desaturation could be accurately predicted from clinical, demographic and dynamic features suggesting the potential for future development of clinical tools for personalised apnoea alarm limits. Given the long-term neurodevelopmental impact of frequent hypoxaemia, understanding infants at greater risk of cardiorespiratory instability is key to improve outcomes for preterm infants.

## Data Availability

All data produced in the present study are available upon reasonable request to the authors.

## Acknowledgements

We would like to thank members of the Paediatric Neuroimaging Group, University of Oxford for assistance with data collection for this project.

## Funding

This work was funded by the Wellcome Trust and Royal Society through a Sir Henry Dale Fellowship awarded to CH (grant reference: 213486/Z/18/Z). YC is funded by the Department of Paediatrics at the University of Oxford and the China Scholarship Council (CSC).

## Author Contributions

YC contributed to conceptualisation, methodology, software, validation, formal analysis, data curation and writing – original draft. VK and SF contributed to investigation and data curation. LBa, FS, CZ, RG contributed to formal analysis. MV and LBe contributed to conceptualisation, formal analysis and supervision. CH contributed to conceptualisation, formal analysis, supervision, project administration, and funding acquisition. All authors contributed to writing – review & editing.

## Competing interests

There are no conflicts of interest.

## Consent statement

Informed written parental consent was obtained prior to the inclusion of each infant in the study.

## Code availability

Code for the statistical analysis and machine learning models is available at: https://gitlab.com/paediatric_neuroimaging/apnoea-bradycardia-desaturation

## Notes

### Competing Interest Statement

The authors have declared no competing interest.

### Author Declarations

The Ethics Committee of the United Kingdom National Research Ethics Service gave ethical approval for this work.

