## Supplementary Material for "Risk of apnoea-related cardiorespiratory instability in preterm infants is modulated by clinical, demographic and dynamic indicators"

**Supplementary Methods**

A. *Machine learning classification to remove false pauses in breathing*

To produce a new machine learning model which accurately identifies true and false short pauses in breathing (5 to 15 seconds), a set of 1248 potential pauses were identified from inter-breath intervals. The potential pauses were manually examined by two independent researchers who rated the pause as either a true pause in breathing or a false positive (for example, caused by shallow breathing or loss of electrode contact). Of these 1248 pauses, 899 were part of a training set and 349 were part of a test set to validate the accuracy of the model. The researchers had an agreement rate of 80%; pauses without consensus were excluded from the training and test sets, and model ratings were compared with researcher ratings to establish accuracy. A machine learning model was trained on the training set using a quadratic Support Vector Machine (SVM) classifier with input variables of the mean absolute amplitude and standard deviation of the IP of the time -10 to -1 seconds relative to the onset of the breathing pause, the period from 1 second after the start of the pause to 1 second before the end of the pause, and the time 1 second to 10 seconds after the end of the pause (similar to the classifier developed for apnoeas > 15 seconds[12]). The support vector machine had a balanced accuracy of 0.93 for the training set and 0.90 for the test set. The confusion matrices for the training and test sets are shown below.

Train set:

| | Predict positive | Predict negative |
|---|---|---|
| Actual positive | 330 | 37 |
| Actual negative | 24 | 508 |

Test set:

| | Predict positive | Predict negative |
|---|---|---|
| Actual positive | 93 | 10 |
| Actual negative | 24 | 222 |

B. *Definition of bradycardia*

To assess the influence of developmental maturity on heart rate responses to apnoea, relationships between postmenstrual age (PMA) and multiple heart rate metrics were examined. For analyses and figures presented in this section, each data point represents, for a given infant, the median across recordings of the within-recording median across all apnoeas. This approach ensures that recordings with a large number of events did not disproportionately influence the results.

Baseline heart rate demonstrated a significant negative correlation with PMA (Pearson correlation coefficient $r = -0.40$, $p < 0.0001$, Supplementary Figure 1A), indicating that older infants exhibited lower resting heart rates. Similarly, the minimum heart rate observed from the onset of an apnoea until 10 s after its termination was also negatively correlated with PMA ($r = -0.33$, $p < 0.0001$, Supplementary Figure 1B). In contrast, the percentage change in heart rate from baseline to the post-apnoeic minimum was not associated with PMA ($r = -0.03$, $p = 0.73$, Supplementary Figure 1C).

These findings highlight substantial maturity-related variation in absolute heart rate values. In particular, baseline heart rate values for late preterm infants were already close to commonly used absolute thresholds for bradycardia. As a consequence, defining bradycardia solely on the basis of a minimum heart rate would potentially classify older infants as experiencing bradycardia, even when the relative deviation from baseline was small and unlikely to reflect clinically meaningful instability. To mitigate this bias and to ensure comparability across developmental stages, bradycardia was therefore defined in this study as a relative decrease in heart rate exceeding 30% from baseline for at least 5 consecutive seconds.

C. Hyperparameter instances examined for the machine learning model (XGBoost)

| **Parameter** | **Instances** |
|---|---|
| Number of estimators (n_estimators) | 200, 400, 800, 1000 |
| Maximum tree depth (max_depth) | 3, 5, 7, 10, 12 |
| Learning rate (learning_rate) | 0.001, 0.01, 0.05, 0.1, 0.2 |
| Subsample ratio (subsample) | 0.5, 0.7, 0.85, 1.0 |
| Column subsample ratio (colsample_bytree) | 0.4, 0.6, 0.8, 1.0 |
| Minimum child weight (min_child_weight) | 1, 3, 5, 7 |
| Minimum loss reduction (gamma) | 0, 0.1, 0.5, 1.0 |
| L1 regularisation (reg_alpha) | 0, 0.01, 0.1, 1.0 |
| L2 regularisation (reg_lambda) | 0.5, 1.0, 2.0, 5.0 |
| Class weight scaling (scale_pos_weight) | 1, 10, 20 |

**Supplementary Figure 1: Relationship between postmenstrual age (PMA) and heart rate characteristics during and after apnoea.** Scatter plots show associations between PMA and (A) baseline heart rate, (B) minimum heart rate observed from the onset of apnoea until 10 s after apnoea termination, and (C) percentage change in heart rate relative to baseline. Each point represents the median value across all apnoeas within an individual recording. Solid red lines indicate the best-fit linear regression.

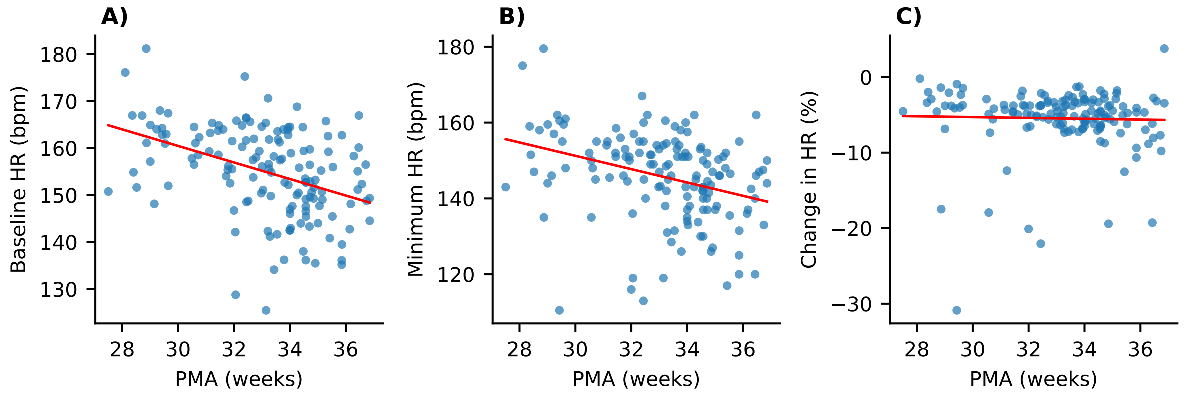

**Supplementary Figure 2: Flowchart of the modelling procedure.** CV = cross-validation. AUROC = area under the receiver operating characteristic curve.

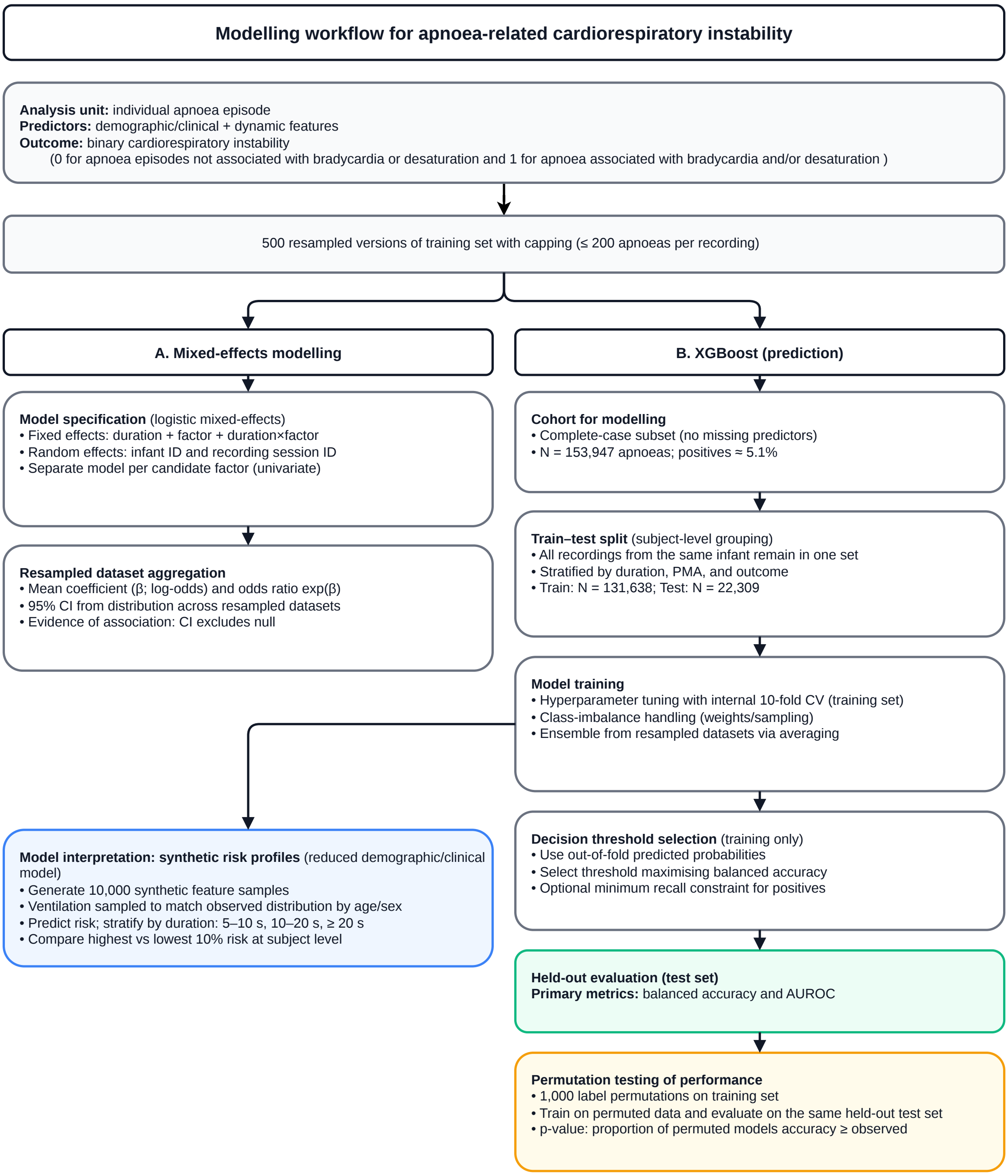

**Supplementary Figure 3: Pairwise heatmaps of the predicted risk of apnoea-related bradycardia and/or desaturation in synthetic data.** Risk was predicted for all apnoeas with durations between 10 and 20 seconds using the optimised XGBoost model. Values shown in each heatmap cell (colour-coded and numerically annotated) represent the mean predicted risk for the corresponding feature combinations.

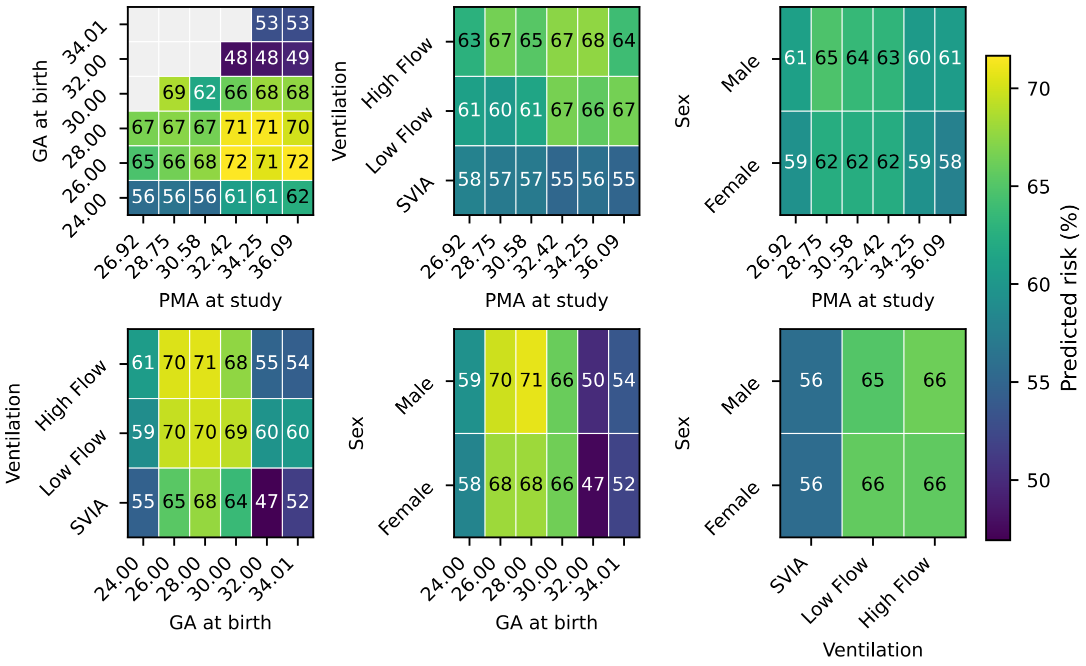

**Supplementary Figure 4: Pairwise heatmaps of the predicted risk of apnoea-related bradycardia and/or desaturation in synthetic data.** Risk was predicted for all apnoeas with durations longer than 20 seconds using the optimised XGBoost model. Values shown in each heatmap cell (colour-coded and numerically annotated) represent the mean predicted risk for the corresponding feature combinations.

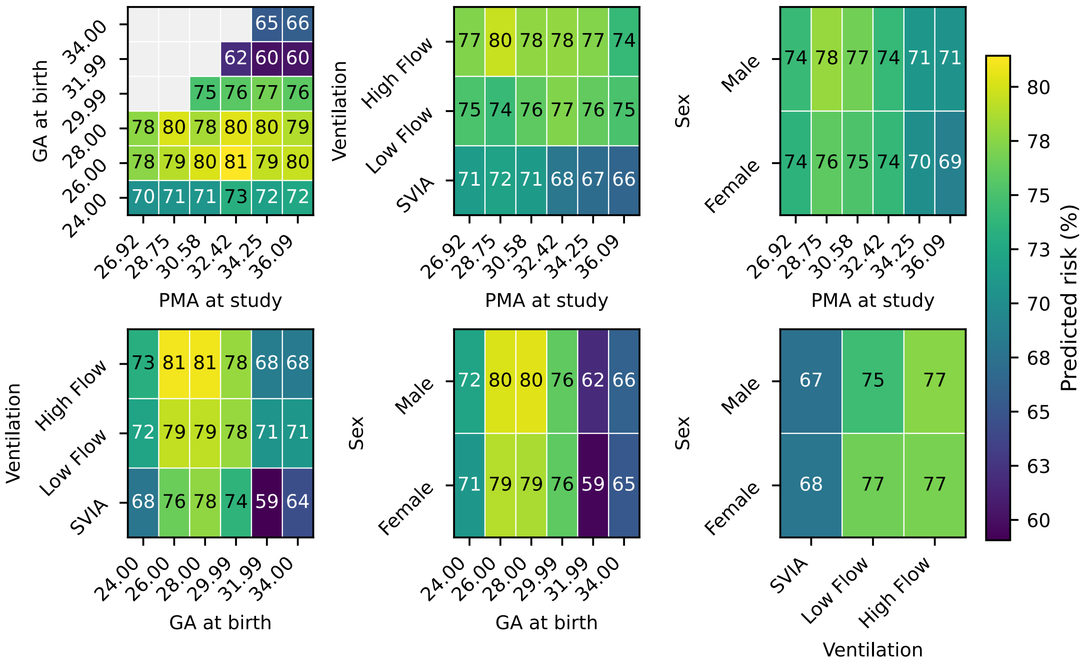

**Supplementary Table 1:** Results from univariate mixed-effects models examining the effects of individual factors and their interactions with apnoea duration on the likelihood that an apnoea is associated with bradycardia and/or desaturation, using only data from recordings less than two weeks long. Significant fraction indicates the percentage of 500 bootstrap replicates in which the effect was significant (see Methods). CI = confidence interval. * significant at the 95 % CI, ** significant at the 99 % CI, *** significant at the 99.9 % CI.

| **Term** | **Significant fraction (%)** | **Mean odds ratio** | **Lower 95% CI** | **Higher 95% CI** | **Significance** |
|---|---|---|---|---|---|
| PMA at study | 100.0 | 1.16 | 1.10 | 1.22 | *** |
| SpO2 baseline | 100.0 | 0.71 | 0.69 | 0.73 | *** |
| Ventilation | 99.6 | 1.29 | 1.16 | 1.45 | *** |
| Weight z-score | 97.6 | 0.84 | 0.76 | 0.91 | *** |
| GA at birth | 89.4 | 0.93 | 0.89 | 0.97 | *** |
| HR baseline | 82.2 | 1.01 | 1.00 | 1.02 | ** |
| Sex | 81.2 | 1.34 | 1.10 | 1.68 | *** |
| Number of apnoeas in last 5 min | 73.6 | 0.94 | 0.89 | 0.98 | ** |
| Apgar (5 min) | 11.2 | 0.98 | 0.92 | 1.05 | |
| Time since last apnoea | 5.4 | 1.00 | 1.00 | 1.00 | |
| Duration:PMA at study | 100.0 | 0.99 | 0.98 | 0.99 | *** |
| Duration:HR baseline | 100.0 | 1.00 | 1.00 | 1.00 | *** |
| Duration:SpO2 baseline | 100.0 | 1.01 | 1.01 | 1.01 | *** |
| Duration:Weight z-score | 96.6 | 1.02 | 1.01 | 1.02 | ** |
| Duration:Sex | 79.4 | 0.98 | 0.95 | 0.99 | ** |
| Duration:Number of apnoeas in last 5 min | 75.4 | 1.01 | 1.00 | 1.01 | * |
| Duration:GA at birth | 42.2 | 1.00 | 0.99 | 1.00 | |
| Duration:Time since last apnoea | 16.8 | 1.00 | 1.00 | 1.00 | |
| Duration:Apgar (5min) | 16.4 | 1.00 | 1.00 | 1.01 | |
| Duration:Ventilation | 13.2 | 1.00 | 0.99 | 1.01 | |

**Supplementary Table 2: Comparison of model features between apnoeas with the highest and lowest 10% predicted risk of bradycardia and/or desaturation in synthetic data.** Features were compared between high- and low-risk groups based on model-predicted risk for all apnoeas with duration between 10 and 20 seconds. Statistically significant differences are indicated as follows: * $p < 0.05$, ** $p < 0.01$, *** $p < 0.001$.

| **Term** | **High-risk mean** | **Low-risk mean** | **p value** | **Significance** |
|---|---|---|---|---|
| PMA at study | 33.58 | 34.31 | < 0.001 | *** |
| GA at birth | 27.65 | 32.15 | < 0.001 | *** |
| Weight z-score | -0.25 | -0.27 | 0.790 | |
| Apgar 5 min | 5.69 | 5.18 | < 0.001 | *** |
| Ventilation==SVIA | 0.15 | 0.90 | < 0.001 | *** |
| Ventilation==Low Flow | 0.18 | 0.03 | < 0.001 | *** |
| Ventilation==High Flow | 0.67 | 0.07 | < 0.001 | *** |
| Sex==female | 0.38 | 0.58 | < 0.001 | *** |
| Sex==male | 0.62 | 0.42 | < 0.001 | *** |

**Supplementary Table 3: Comparison of model features between apnoeas with the highest and lowest 10% predicted risk of bradycardia and/or desaturation in synthetic data.** Features were compared between high- and low-risk groups based on model-predicted risk for all apnoeas with duration longer than 20 seconds. Statistically significant differences are indicated as follows: * $p < 0.05$, ** $p < 0.01$, *** $p < 0.001$.

| Term | High-risk mean | Low-risk mean | p value | Significance |
|---|---|---|---|---|
| PMA at study | 32.41 | 34.78 | < 0.001 | *** |
| GA at birth | 27.39 | 32.45 | < 0.001 | *** |
| Weight z-score | -0.48 | -0.18 | < 0.001 | *** |
| Apgar 5 min | 6.31 | 4.68 | < 0.001 | *** |
| Ventilation==SVIA | 0.06 | 0.96 | < 0.001 | *** |
| Ventilation==Low Flow | 0.13 | 0.02 | < 0.001 | *** |
| Ventilation==High Flow | 0.81 | 0.02 | < 0.001 | *** |
| Sex==female | 0.39 | 0.52 | < 0.001 | *** |
| Sex==male | 0.61 | 0.48 | < 0.001 | *** |
